# Comparing a new device to measure lung volumes with three traditional methods in individuals with different lung conditions

**DOI:** 10.1101/2021.03.05.21252831

**Authors:** Jacob Zac, Salomon Zac, Rogelio Pérez-Padilla, Arantxa Remigio-Luna, Nicolas Guzmán-Boulloud, Laura Gochicoa-Rangel, Carlos Guzmán-Valderrábano, Ireri Thirión-Romero

## Abstract

**Background:** Lung volumes can be measured by body plethysmography (BP), by inert gas dilution during a single-breath or multiple breaths and by radiographic methods based on chest roentgenogram or CT scanning. Our objective was to analyze the concordance between several methods including a new instrument in a variety of pulmonary conditions.

**Methods:** We recruited four groups of adult volunteers at the COPD and Tobacco Clinic of a respiratory referral hospital: patients with lung bullae, with obstructive lung diseases, with restrictive lung diseases and healthy controls; all subjects underwent lung volume measurements according to ATS/ERS standards in random order with each method and then CT scanning. Differences among groups were estimated by Kruskal-Wallis tests. Concordance correlation coefficients (CCC) and Bland-Altman plots were performed.

**Results:** Sixty-two patients were studied including 15 with lung bullae, 14 with obstructive lung diseases, 12 with restrictive lung disease and 21 healthy subjects. Highest concordance was obtained between BP and CT scanning (CCC 0.95, mean difference −0.35 L) and the lowest, with TLC-DLCO_sb_ (CCC 0.65, difference - 1.05 L). TLC measured by BP had a moderate concordance with Minibox (CCC=0.91, mean difference −0.19 L). Minibox, on the other hand had the lowest intratest repeatibility (2.7%) of all tested methods.

**Conclusions:** Lung volumes measured by BP and CT had a substantial concordance in the scenario of varied pulmonary conditions including lung bullae, restrictive and obstructive diseases. The new minibox device, had low intratest variability, and was easy to perform, with a reasonable concordance with BP.

## Introduction

The measurement of lung volumes, essential to identify abnormalities such as lung restriction, can be performed through different techniques including lung function tests and thoracic imaging.^1^ Body plethysmography (BP), the best studied method, measures ventilated and non-ventilated intrathoracic gas volume (ITGV) based on Boyle’s law, and requires that the subject be inside a closed cabin during the maneuver.^2,3^. However, the device is expensive and large in size making it hardly accessible, especially in the office setting and, in addition, the quality control is complex. For patients, it can be intimidating, uncomfortable and the maneuvers a challenge to perform, especially in the presence of certain conditions such as claustrophobia or advanced lung disease.^3^

Measuring ventilated lung volume with a tracer gas, such as that estimated during single breath diffusing lung capacity for carbon monoxide single breath (DLCO_sb_), is simple and widely used method for evaluation of lung function status, but it tends to underestimate intrathoracic gas volume in the presence of airflow obstruction, maldistribution of ventilation, or lung bullae.^4^

Currently it is also possible to measure pulmonary volumes with imaging methods such as computed tomography (CT). In recent years, various tools have been developed that have improved the quantitative analysis of CT and have been studied for pre-operative assessment, to evaluate therapeutic interventions,^5–9^ and to define repeatability.^10,11^ However, this method is expensive and involves exposure to radiation; thus, CT has not proven its cost-effectiveness compared with the conventional pulmonary function test (PFT).^3^

The MiniBox+™ is a new cabin-less compact device, claiming to measure rapidly and accurately absolute lung volumes in subjects with either normal, obstructive or restrictive lung status with minimal cooperation from subjects.^12–14^

Our objective is to compare lung volume measurements (LVM) by different methods, such as BP, DLCO_sb_, CT and the new cabin-less plethysmography in patients with varied lung volumes and conditions, as well as in healthy subjects.

## Methods

The study, approved by the Institutional Ethics Committee (Code C63-18), was conducted at the Department of Research on Smoking and COPD at the Instituto Nacional de Enfermedades Respiratorias (INER) Ismael Cosío Villegas, a referral center for respiratory diseases mainly for uninsured patients in Mexico City, at 2,240 m above sea level. All participants signed an informed consent document.

Between October 2018 and May 2019 we prospectively identified and recruited adult patients who attended the clinic who met study inclusion criteria and healthy subjects who wanted to participate in the research. We considered a total of four groups with conditions often requiring lung volume analysis, and with a wide range of lung volumes: patients with airflow obstruction, patients with lung bullae both with known difficulty for lung volume measurements by inert gas dilution methods, also a group with lung restriction and subjects healthy from the respiratory point of view.

The group with airflow obstruction, defined as post-bronchodilator (Albuterol 400 µg) spirometry with FEV_1_/FVC <lower limit of normality (LLN), had all a clinical diagnosis of chronic obstructive pulmonary disease (COPD).

Another group of patients had lung bullae by chest CT scanning, with or without airflow obstruction, and had a separate outpatient clinic and follow up, aimed to identify candidates for bullectomy or invasive procedures.

The group with restrictive lung disease had a post-bronchodilator spirometry with normal FEV_1_/FVC, FVC <80% predicted value, and a TLC <80% of predicted, and several clinical diagnosis including interstitial pulmonary disease due to varied causes such as hypersensitivity pneumonitis, connective tissue disease, or with obesity-hypoventilation syndrome.

In the group of healthy subjects all were volunteers with normal spirometry, never smokers (<100 smoked cigarettes in a lifetime), lacking a personal history of pulmonary, cardiac, renal, hepatic, or metabolic diseases, or respiratory symptoms in the last 4 weeks or persistent respiratory symptoms (>3 days/week) in the previous 12 months. All the volunteers were employees of the institute or relatives of the patients who accepted participate.

We excluded pregnant women, patients with acute respiratory disease or exacerbation, and those admitted to hospital within the last 4 weeks.

After assisting at an explanation of the study in detail, the subjects read and signed informed consent, answered the modified Medical Research Council dyspnea scale (mMRC), and the Short Form 12 (SF-12) health-related quality of life instrument. We also documented exacerbations in the past 12 months, use of supplemental oxygen, and the presence of comorbidities. Subsequently, weight and height were measured using a *SECA*^*®*^ digital scale and stadiometer (models mBCA 514 and 274, respectively).

Simple helical CT (Siemens Somatom Definition A 128) was the first study performed. The mediastinal window images were acquired with 110 mA, 120 kV, Pitch 1.4, FOV 362, and reconstructed with 3mm thickness, utilizing the algorithm for mediastinum I26 homogeneous. For the lung window, acquisition was with 110 mA, 120 kV, Pitch 1.4, FOV 362, and reconstructed with 1 mm thickness, with a lung algorithm I70f very defined. We explained in detail to the patient how to perform a TLC maneuver and how to practice it before conducting the study; then the patient was instructed by means of a simple video. Subsequently, Osirix-Lite software (V10.10) was used to estimate lung volume, through semi-automatic segmentation of pulmonary fields, with density thresholds of −1,024 to −200 Hownsfield units. No correction was made for any study, and volume includes anatomic dead space. CT was performed in all patients except for healthy subjects.

The PFT were performed in the morning within a period of up to 3 h, at the same laboratory and by the same two technicians. None of the subjects used bronchodilators 24 h prior to performing the tests. Before testing, the maneuvers were explained and demonstrated. All tests were performed according to ATS/ERS standards.^3,4,15,16^ Flow calibration was verified daily with a certified 3-L syringe, flow linearity was tested weekly at three different flow rates, considering as acceptable a variation of ±2.5%. Ambient conditions (barometric pressure, temperature and humidity) were updated automatically and cabin’s pressure transducer calibration was performed daily. The DLCO_sb_ was performed with the following concentration of gases: Helium 10.4 cmol/mol; CO 0.313 cmol/mol, and O_2_ 21.69 cmol/mol, balanced with nitrogen.

The spirometry (Easy One Pro Lab, Ndd®, Zurich, Switzerland, and PulmoOne Advance Medical Devices, Ltd., Ra’anana, Israel) was carried out with a nasal clip and with the subject seated. Individuals took a quick full inspiration followed by an explosive forced expiration. At least three technically acceptable maneuvers were performed, with two best FVC and FEV_1_ within 150 mL to fulfill repeatability criteria. BP (Masterscreen Body, Lab Manager, CareFusion, Germany) was performed with the subject seated inside the closed cabin, using a nasal clip and holding hands against cheeks. The subject then began to take tidal breaths until stabilization. When told, the subject performed a panting maneuver against a closed shutter, to measure ITGV (at functional residual capacity) with a frequency of >0.5-<1.5 Hz, followed by an inspiratory capacity maneuver, and finally, slow vital capacity (SVC). At least three technically acceptable maneuvers were performed, with an ITGV variance of <5% and vital capacity (VC) difference of <0.15L to ensure test repeatability.

Measurement of TLC by cabin-less plethismography (PulmoOne Advance Medical Devices, Ltd. Ra’anana, Israel) was carried out with a nasal clip and hands held against cheeks; then, after reaching stable tidal breathing, the patient performed an inspiratory capacity maneuver and finally, a slow vital capacity maneuver. During tidal breathing, in the inspiratory phase, several transient airway occlusions occur in which airflow to and from the reservoir and pressure, are measured. These measurements are utilized for estimation of total lung capacity by regression equations obtained from a group of healthy individuals as well from patients with several diseases.^17^ A maneuver was considered acceptable when tidal breathing was stable and at least six occlusions took place. Good repeatability criteria was considered as at least three technically satisfactory maneuvers with an ITGV of <5% variance, a VC difference of <0.15 L between the two highest, and a TLC of <0.20L between the two highest.

The DLCO_sb_ (Easy One Pro Lab (Ndd®, Zurich, Switzerland) was performed with the subject seated with a nasal clip. The participant performed 4-6 tidal breaths until stabilization; then the participant was asked to expire maximally, followed by maximal quick inspiration (of the test gas) with 10-s breath-holding and then a full exhalation. The time between procedures was at least 4 min. At least two technically acceptable maneuvers of A or B grade quality were completed, in order to reach two DLCO_sb_ within 2 mL/min/mmHg of each other for repeatability criteria. Reference values used for this study were NHANES III for spirometry^18^, Quanjer’s for plethysmography^19^ and Vazquez’s for DLCO ^20^

### Statistical analysis

PFT sequence was performed in random order (Research Randomizer, Version 4.0) (http://www.randomizer.org/). Comparison among groups was made using the Kruskal-Wallis for continuous variables and Fisher exact tests for nominal or dichotomic variables. To determine agreement among TLC in different groups and methods, we utilized Bland-Altman plots and the Lin’s concordance correlation coefficient (CCC). We calculated the coefficient of variation (CV, standard deviation/mean) to compare intra-test variability among devices. P value was considered statistically significant if <0.05. To perform all statistical analyses, we employed STATA Ver. 13 statistical software on a Macintosh system (StataCorp, Collage Station, TX, USA).

## Results

A total of 69 patients were recruited, but seven were eliminated, five because of poor quality criteria in pulmonary function tests and two because airflow obstruction disappeared in the post-bronchodilator test. Sixty-two subjects were analyzed, including 21 healthy subjects, 15 with lung bullae, 14 with obstructive lung disease, and 12 with restrictive lung disease.

The general characteristics of the population are presented in **Table 1**. Women predominated in the healthy and restrictive lung groups. The healthy group was the youngest, and the obstructive lung group, the oldest subjects. Body mass index (BMI) in the restrictive lung group was highest among the groups (27 kg/m^2^).

**Table 1.** Baseline characteristics of the population

|  | **Total**<br>**62** | **Healthy**<br>**21** | **Obstructive**<br>**14** | **Bullae**<br>**15** | **Restrictive**<br>**12** | **P\*** |
| --- | --- | --- | --- | --- | --- | --- |
| **Women** | 40<br>(61%) | 16<br>(76%) | 3<br>(21%) | 7<br>(47%) | 11<br>(92%) | <0.001 |
| **Age**<br>**(years)** | 55<br>(28-67) | 25<br>(24-29) | 67<br>(60-68) | 56<br>(52-71) | 59<br>(54-71) | <0.001 |
| **Height (cm)** | 162<br>(154-167) | 162<br>(159-165) | 161<br>(158-166) | 167<br>(156-171) | 151<br>(142-161) | <0.01 |
| **Weight (kg)** | 63<br>(56-77) | 60<br>(57-69) | 65<br>(55-79) | 69<br>(56-81) | 67<br>(52-78) | 0.94 |
| **BMI (kg/m<sup>2</sup>)** | 25<br>(21-30) | 21<br>(20-24) | 24<br>(21-31) | 25<br>(20-33) | 27<br>(25-31) | 0.02 |
| **FEV<sub>1</sub>/FVC** | 79<br>(62-85) | 83<br>(79-86) | 58<br>(36-62) | 70<br>(46-79) | 86<br>(82-87) | <0.001 |
| **FEV<sub>1</sub>%p** | 73 (51-92) | 98<br>(87-106) | 54<br>(36-78) | 54<br>(28-77) | 60<br>(53-72) | <0.001 |
| **TLC%p** | 104<br>(82-114) | 111<br>(103-117) | 121<br>(110-131) | 96<br>(76-111) | 61<br>(57-66) | <0.001 |
| **Exacerbations last**<br>**year** | 25<br>(38%) | 0 | 10<br>(71%) | 8<br>(53%) | 7<br>(44%) | <0.001 |
| **Pack-years of**<br>**smoking** | 28<br>(8-43) | 0 | 40<br>(32-49) | 15<br>(8-25) | 1<br>(1-27) | 0.002 |
| **Hour-years of**<br>**exposure to**<br>**biomass stove** | 36<br>(21-54) | 0 | 40<br>(32-48) | 90<br>(30-104) | 30<br>(12-42) | 0.23 |
| **mMRC dyspnea**<br>**score** | 1<br>(0-2) | 0 | 1<br>(1-2) | 2<br>(1-3) | 2<br>(2-3) | <0.001 |

Variables are expressed in medians, p25-p75, or frequencies according to type. *P*\* was calculated with Kruskal-Wallis and Fisher exact tests, according to the type of variable. BMI: Body mass index, CAT: mMRC.

Restrictive lung group had a TLC expressed as percentage of predicted (TLC%p) of 61(57-66) measured by BP; patients with obstructive lung disease had a post bronchodilator FEV_1_%p of 54 (36-78). The obstructive group had the highest number of exacerbations (71%) and highest smoking intensity (40 pack-years). Home oxygen therapy was more common in the restrictive lung group.

TLC measured by BP ranged between 3.8 L and 6.6 L in the participants, and the ITGV, between 2.2 L and 4.3 L. We analyzed the agreement of TLC measured by BP (our gold standard) and the other methods in all groups as presented in **Table 2**, including the CCC, the intercept (alpha α) and slope (beta β) of a linear regression. In a perfect concordance, CCC would be 1, with α=0 and β=1. Best agreement was observed between BP and CT (CCC=0.95, α −0.18, β 0.97, and mean difference, −0.35L). Agreement between BP and DLCO_sb_ was significantly lower with CCC=0.65 and a large average difference of −1.05L, while BP vs. Minibox+™ was moderate (CCC=0.91, with a mean difference of −0.19 L).

**Table 2.** TLC degree of agreement between BP and the other devices in all participants, and linear regression estimates.

| Device | Cabin-less plethysmography | DLCO<sub>sb</sub> | CT scan |
| --- | --- | --- | --- |
| **CCC**<br>(BP vs. all other methods) | 0.91 | 0.65 | 0.95 |
| **Δ (L)** | -0.19±0.75 | -1.05±1.41 | -0.35±0.55 |
| **95% LA** | -1.67 to 1.28 | -3.29 to 1.19 | -1.44 to 0.74 |
| **α** | 0.45 | -0.21 | -0.18 |
| **β** | 0.88 | 0.84 | 0.97 |

Agreement was calculated with CCC: concordance correlation coefficient, α: intercept; β: slope, Δ: mean difference average in L, 95% LA: 95% limits of agreement.

In **Table 3**, we observe the agreement of TLC measured by BP and by the other methods studied separated by groups. Concordance in restrictive subjects between BP and cabin-less plethysmography was highest comparing with the other groups (CCC=0.90), following by healthy and bullae lung groups (CCC=0.87 and CCC=0.85, respectively). Concordance with DLCO_sb_ in the obstructive group and in patients with bullae were the lowest (CCC=0.18 and 0.47, respectively); also for healthy and restrictive groups the concordance was < 0.90. Volumes measured by CT scan had highest agreement with BP for lung bullae and obstructive groups (CCC 0.95 and CCC 0.76, respectively).

**Table 3.**
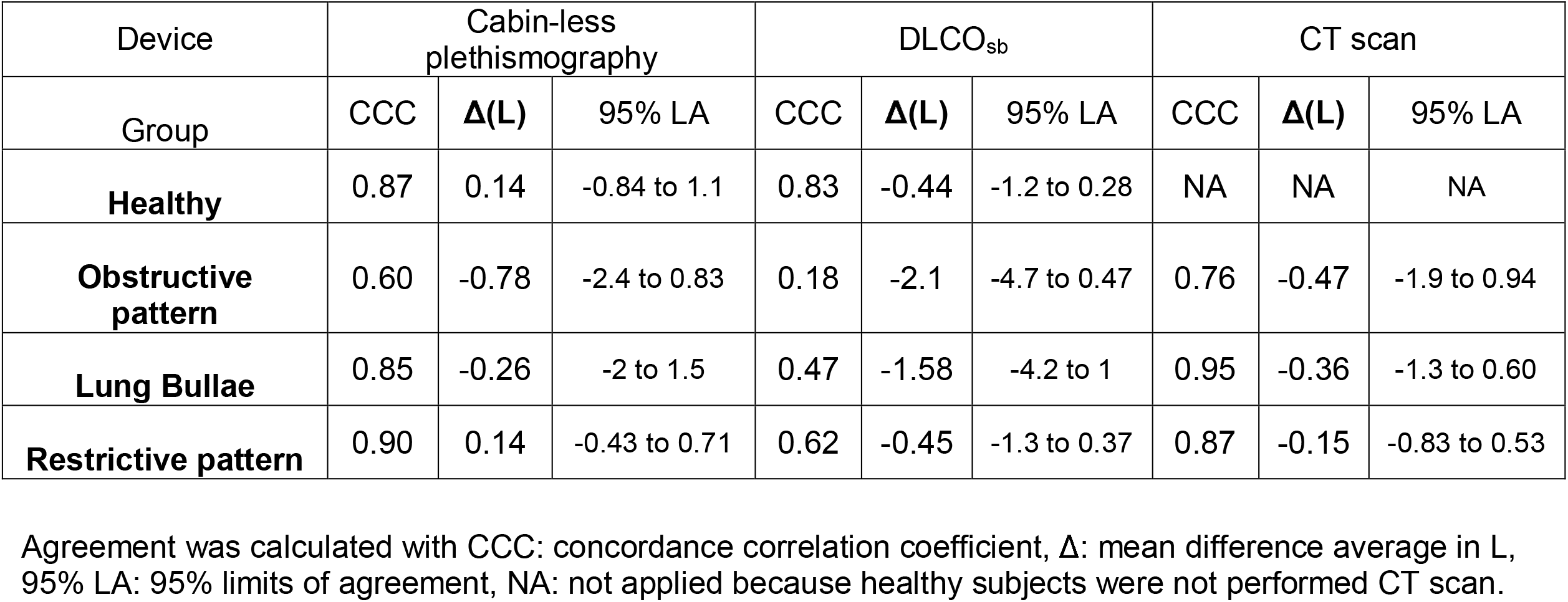
Subgroup agreement analysis of TLC between BP and other devices

Agreement is shown graphically in Bland-Altman plots (**Figure 1**). The method with highest agreement compared with BP was chest CT, then with cabin-less plethysmography following close behind, and DLCO_sb_ with higher discrepancy, especially in patients with obstructive and lung bullae groups.

**Figure 1.**
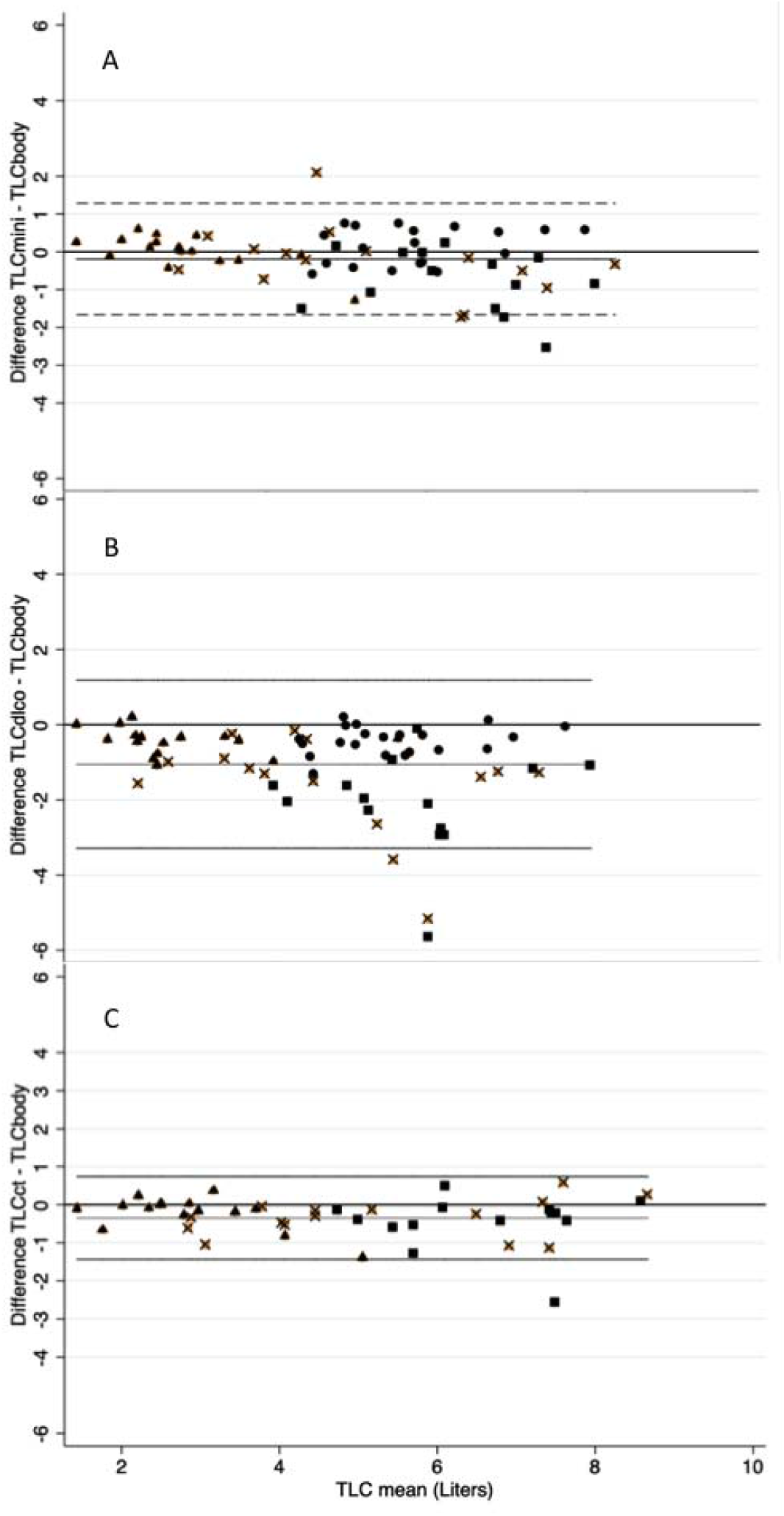
Bland-Altman plots shows **Panel A)** TLCbody vs. TLCmini; **Panel B**) TLCbody vs. TLCdlco; **Panel C)** TLCbody vs. TLCct. Subjects groups: · healthy, X lung bullae, ▪ obstructive disease, Δ restrictive disease.

The agreement of VC, ITGV, and RV in all subjects between cabin-less plethismography and BP was 0.85, 0.81, and 0.76, respectively. We calculated the intratest-CV and compared this among devices (except by CT scanning, performed only once). For TLC, the CV of cabin-less plethismography was the lowest with 2.7%, followed by BP (3.3%) and DLCO_sb_ (4.6%).

Time spent in performing each method had a mean of 12.5 min (SD 3.4) for cabin-less plethismography, 19.2 (SD 9.6) for BP, and 12.6 (SD 6.3) for DLCO_sb_. Participants were asked about the difficulty they encountered in to performing each test, with five levels (1-very hard to 5-very easy); 76% considered cabin-less plethismography a test that can be carried out between easily and very easily; measuring lung volumes by BP was considered easy and very easy by 35% of the participants.

## Discussion

We compared several methods of lung volume measurement (LVM) in different groups of participants with a wide variety of lung volumes and health conditions, including a new method: Minibox™. ATS/ERS guidelines describe several methods for LVM but for the purposes of this work, we considered BP the gold standard.^3^ Our results demonstrated that the best agreement observed of TLC was between BP and CT scan with a substantial concordance (CCC=0.95) including among individuals in the lung bullae group, as found in previous studies^21–25^, whereas agreement was moderate between BP and cabin-less plethysmography (CCC=0.91) in the whole group of individuals tested. We also identified variations in the concordance for the different participant groups and volumes measured.

Although CT could represent an accurate and reliable method to measure lung volumes, requires a breath-hold in maximal inspiration (TLC) during the study that could be challenging to perform in subjects with advanced lung disease. In addition, measurement is costly, depends on the respiratory cycle and the presence of pathologic conditions,^26^, exposes the patients to radiation, and segmenting the lung fields is difficult. Nevertheless, it may be cost-effective if a CT ordered for other reasons, is utilized to measure lung volumes.

In healthy subjects the concordance between BP and cabin-less plethysmography was 0.87. Agreement of BP and DLCO_sb_ was 0.83. In this comparison we lacked the analysis with CT scan as was not performed in healthy subjects; in the case of restrictive group cabin-less plethysmography was the best method with a moderate concordance with BP (CCC=0.90), followed by CT scan with a CCC=0.87 (considered poor) and DLCO_sb_ with a low concordance (CCC=0.62). Other studies which included restrictive patients and healthy subjects had reported a concordance up to 0.81.^21,27^

Among subjects with COPD, we found poor agreement (CCC=0.60) between BP and cabin-less plethysmography, with an underestimation of TLC by cabin-less plethysmography. Despite this, in the lung bullae group, which was challenging because of the presence of unventilated air spaces, agreement between BP and cabin-less plethysmography was better (CCC=0.85), demonstrating a reasonable measurement of intrathoracic gas, but one that is affected by airflow obstruction. We must also consider the reported systematic overestimation of lung volumes by plethysmography in the presence of severe airflow obstruction due to poor equilibration between alveolar and mouth pressure during the panting maneuver, with the rate of overestimation proportional to the FEV_1_. However, in nearly all studies, this overestimation was corrected by performing this maneuver within a panting frequency in a range between >0.5 - <1Hz.^28–32^

The agreement between BP and DLCO_sb_ was poor in the presence of lung bullae and airflow obstruction (CCC=0.47 and CCC=0.18, respectively), consistent with other studies, finding that gas dilution method by a single breath, only measures correctly well ventilated lung volume.^19,27,33–37^

In a previous study including measurement of lung volumes with different methods^34^, a systematic overestimation of TLC with BP was observed, when compared with DLCO rebreathing (DLCO_rb_) (Δ0.63L) and CT (Δ1.08), despite adherence to proper technique. Likely the main reason for these discrepancies may be explained by the use of rebreathing DLCO instead of DLCO_sb_ done in a single breath, and the exclusion of the anatomical death space in CT^38,39^. Also, in our study the measuring of TLC with CT includes the anatomical dead space.

We found that cabin-less plethysmography had the lowest intra-test CV in TLC (2.7%), the shortest time for the test and was perceived as easier to perform than BP and DLCO_sb_ regardless of the lung status. Both Cohen et al, and Fredberg, et al ^12–14^ reported that BP and cabin-less method measured similar TLC in obstructive patients, but in our study, even though the cabin-less plethysmography performed well in healthy and restrictive subjects, in subjects with obstruction and pulmonary bullae both methods had a poor agreement. The difference found between the BP and cabin-less plethysmography (−0.19±0.75 L) or between BP and CT scan (−0.35±0.55) have to be taken into account for interpretation. Do not consider the different methods to measure lung volume as interchangeable.

Our study had several limitations. First, we only performed the LVM once (including several maneuvers except for CT scanning, only one study); therefore, we could not analyze the inter-test variability and evaluate its usefulness as a longitudinal assessment.^10,11^ Second, the group of healthy subjects was younger compared with the other groups and lacked CT measurement. Third, we performed the CT measurement without a spirometric control, as did Tantucci and colleagues^21^, the best way to ensure that individuals were tested at TLC, but CT is performed reasonably well at TLC under routine conditions, and spirometric control of TLC is not available in the majority of centers. Last, our studied population was small, especially when the data is analyzed by group.

## Data Availability

Data and Code AvailabilityThe data and codes related to the findings of this study will be available from the corresponding author after publication upon reason-able request. The research team will provide an email address for communication once the data are approved to be shared withothers. The proposal with detailed aims, statistical plan, and other information/materials may be required to guarantee the rationalityof requirement and the security of the data. The patient-level data, but without names and other identifiers, will be shared after reviewand approval of the submitted proposal and any related requested materials.

## Conclusions

The cabin-less plethysmography proved to be an easier and faster method than any other presented here. Lung volumes measured by BP had a substantial concordance with those measured by CT scanning, and moderate with the Minibox. A poor concordance was seen when lung volumes were measured by DLCO_sb_ in individuals with bullae or airflow obstruction.

## ACKNOWLEDGMENTS

The Minibox device was lent by the distributor in Mexico at no cost. The company did not offer any additional grant or help and did not participate in the design, of the study, recruitment, analysis of results, or in the writing of the manuscript.

We thank all participants of the study.

## References

1. Luo J, Liu D, Chen G, Liang B, Liu C. Clinical Roles of Lung Volumes Detected by Body Plethysmography and Helium Dilution in Asthmatic Patients: A Correlation and Diagnosis Analysis. Sci Rep. 2017;7(1):40870. doi:10.1038/srep40870

2. Dubois AB, Botelho SY, Bedell GN, Marshall R, Comroe JH. A rapid plethysmographic method for measuring thoracic gas volume: a comparison with a nitrogen washout method for measuring functional residual capacity in normal subjects. J Clin Invest. 1956;35(3):322–326. doi:10.1172/JCI103281

3. Wanger J, Clausen JL, Coates A, et al. Standardisation of the measurement of lung volumes. Eur Respir J. 2005;26(3):511–522. doi:10.1183/09031936.05.00035005

4. Graham BL, Brusasco V, Burgos F, et al. 2017 ERS/ATS standards for single-breath carbon monoxide uptake in the lung. Eur Respir J. 2017;49(1):1600016. doi:10.1183/13993003.00016-2016

5. Strange C, Herth FJ, Kovitz KL, et al. Design of the Endobronchial Valve for Emphysema Palliation Trial (VENT): a non-surgical method of lung volume reduction. BMC Pulm Med. 2007;7(1):10. doi:10.1186/1471-2466-7-10

6. Gierada DS, Yusen RD, Pilgram TK, et al. Repeatability of Quantitative CT Indexes of Emphysema in Patients Evaluated for Lung Volume Reduction Surgery. Radiology. 2001;220(2):448–454. doi:10.1148/radiology.220.2.r01au46448

7. Coxson HO, Nasute Fauerbach PV., Storness-Bliss C, et al. Computed tomography assessment of lung volume changes after bronchial valve treatment. Eur Respir J. 2008;32(6):1443–1450. doi:10.1183/09031936.00056008

8. Ostridge K, Wilkinson TMA. Present and future utility of computed tomography scanning in the assessment and management of COPD. Eur Respir J. 2016;48(1):216–228. doi:10.1183/13993003.00041-2016

9. Muller NL, Coxson H. Chronic obstructive pulmonary disease * 4: Imaging the lungs in patients with chronic obstructive pulmonary disease. Thorax. 2002;57(11):982–985. doi:10.1136/thorax.57.11.982

10. Brown MS, Kim HJ, Abtin F, et al. Reproducibility of Lung and Lobar Volume Measurements Using Computed Tomography. Acad Radiol. 2010;17(3):316–322. doi:10.1016/j.acra.2009.10.005

11. Shin JM, Kim TH, Haam S, et al. The repeatability of computed tomography lung volume measurements: Comparisons in healthy subjects, patients with obstructive lung disease, and patients with restrictive lung disease. Nolan A, ed. PLoS One. 2017;12(8):e0182849. doi:10.1371/journal.pone.0182849

12. Adam O, Shiner R, Calverley P, Solway J, Brown R, Fredberg J. The measurement of absolute lung volume without plethysmography. Eur Respir J. 2011;38(Suppl 55).

13. Fredberg JJ, Brown R, Adam O, et al. Absolute Lung Volumes In Healthy Subjects By The Partial Volume Method. In: American Thoracic Society International Conference Meetings Abstracts. American Thoracic Society; 2012:A5572–A5572. doi:10.1164/ajrccm-conference.2012.185.1_meetingabstracts.a5572

14. Cohen I, LaPrad A, Adam O, et al. Determination of total lung capacity (TLC) without body plethysmography. Eur Respir J. 2013;42(Suppl 57).

15. Graham BL, Steenbruggen I, Barjaktarevic IZ, et al. Standardization of spirometry 2019 update an official American Thoracic Society and European Respiratory Society technical statement. Am J Respir Crit Care Med. 2019;200(8):E70–E88. doi:10.1164/rccm.201908-1590ST

16. Miller MR, Hankinson J, Brusasco V, et al. Standardisation of spirometry. Eur Respir J. 2005;26(2):319–338. doi:10.1183/09031936.05.00034805

17. Adam O, Cohen I, Yip WK, et al. Total lung capacity without plethysmography. bioRxiv. 2018:1–22. doi:10.1101/395160

18. Odencrantz OHNR, Fedan KB. Spirometric Reference Values from a Sample of the General U.S. Population. 1994.

19. Stocks J, Quanjer PH. Reference values for residual volume, functional residual capacity and total lung capacity. ATS Workshop on Lung Volume Measurements. Official Statement of The European Respiratory Society. Eur Respir J. 1995;8(3):492–506.

20. Vázquez-García JC, Pérez-Padilla R, Casas A, et al. Reference values for the diffusing capacity determined by the single-breath technique at different altitudes: The Latin American single-breath diffusing capacity reference project. Respir Care. 2016;61(9):1217–1223. doi:10.4187/respcare.04590

21. Tantucci C, Bottone D, Borghesi A, Guerini M, Quadri F, Pini L. Methods for Measuring Lung Volumes: Is There a Better One? Respiration. 2016;91(4):273–280. doi:10.1159/000444418

22. Garfield JL, Marchetti N, Gaughan JP, Steiner RM, Criner GJ. Total lung capacity by plethysmography and high-resolution computed tomography in COPD. Int J Chron Obstruct Pulmon Dis. 2012;7:119–126. doi:10.2147/COPD.S26419

23. Zaporozhan J, Ley S, Eberhardt R, et al. Paired Inspiratory/Expiratory Volumetric Thin-Slice CT Scan for Emphysema Analysis. Chest. 2005;128(5):3212–3220. doi:10.1378/chest.128.5.3212

24. Gierada DS, Hakimian S, Slone RM, Yusen RD. MR analysis of lung volume and thoracic dimensions in patients with emphysema before and after lung volume reduction surgery. Am J Roentgenol. 1998;170(3):707–714. doi:10.2214/ajr.170.3.9490958

25. Jung WS, Haam S, Shin JM, et al. The feasibility of CT lung volume as a surrogate marker of donor–recipient size matching in lung transplantation. Medicine (Baltimore). 2016;95(27):e3957. doi:10.1097/MD.0000000000003957

26. Mansoor A, Bagci U, Foster B, et al. Segmentation and Image Analysis of Abnormal Lungs at CT: Current Approaches, Challenges, and Future Trends. RadioGraphics. 2015;35(4):1056–1076. doi:10.1148/rg.2015140232

27. Tang Y, Zhang M, Feng Y, Liang B. The measurement of lung volumes using body plethysmography and helium dilution methods in COPD patients: a correlation and diagnosis analysis. Sci Rep. 2016;6(1):37550. doi:10.1038/srep37550

28. Shore SA, Huk O, Mannix S, Martin JG. Effect of Panting Frequency on the Plethysmographic Determination of Thoracic Gas Volume in Chronic Obstructive Pulmonary Disease 18211;3. Am Rev Respir Dis. 1983;128(1):54–59. doi:10.1164/arrd.1983.128.1.54

29. Rodenstein DO, Stănescu DC. Frequency dependence of plethysmographic volume in healthy and asthmatic subjects. J Appl Physiol. 1983;54(1):159–165. doi:10.1152/jappl.1983.54.1.159

30. Ruppel GL, McCarthy K, Siu A, Stoller JK. What is the clinical value of lung volumes? Respir Care. 2012;57(1):26–35; discussion 35-8. doi:10.4187/respcare.01374

31. Stanescu DC, Rodenstein D, Cauberghs M, Van de Woestijne KP. Failure of body plethysmography in bronchial asthma. J Appl Physiol. 1982;52(4):939–948. doi:10.1152/jappl.1982.52.4.939

32. Shinozaki S, Matsuzawa Y, Yoshikawa S, et al. [Effect of natural panting frequency and cheek support on the plethysmographic determination of thoracic gas volume in patients with chronic obstructive pulmonary disease]. Nihon Kyobu Shikkan Gakkai Zasshi. 1991;29(8):954–962.

33. Vestbo J, Anderson W, Coxson HO, et al. Evaluation of COPD Longitudinally to Identify Predictive Surrogate End-points (ECLIPSE). Eur Respir J. 2008;31(4):869–873. doi:10.1183/09031936.00111707

34. O’Donnell CR, Bankier AA, Stiebellehner L, Reilly JJ, Brown R, Loring SH. Comparison of Plethysmographic and Helium Dilution Lung Volumes: Which Is Best for COPD? Chest. 2010;137(5):1108–1115. doi:10.1378/CHEST.09-1504

35. Coates AL, Peslin R, Rodenstein D, Stocks J. Measurement of lung volumes by plethysmography. Eur Respir J. 1997;10(6):1415–1427.

36. Milite F, Lederer DJ, Weingarten JA, Fani P, Mooney AM, Basner RC. Quantification of single-breath underestimation of lung volume in emphysema. Respir Physiol Neurobiol. 2009;165(2-3):215–220. doi:10.1016/j.resp.2008.12.009

37. Cazzola M, Rogliani P, Curradi G, et al. A pilot comparison of helium dilution and plethysmographic lung volumes to assess the impact of a long-acting bronchodilator on lung hyperinflation in COPD. Pulm Pharmacol Ther. 2009;22(6):522–525. doi:10.1016/j.pupt.2009.05.005

38. Stanescu DC. How To Measure Lung Volume? Chest. 2010;138(5):1280–1281. doi:10.1378/CHEST.10-1369

39. Tantucci C. Comparison Among Different Methods on Measurement of Total Lung Capacity in COPD. Chest. 2010;138(2):458–459. doi:10.1378/CHEST.10-0369

